# Trends in childhood hospital admissions for carious tooth extractions in England in relation to the UK soft drink industry levy: an interrupted time series analysis of Hospital Episode Statistics

**DOI:** 10.1101/2023.02.27.23286504

**Authors:** Nina T. Rogers, David I. Conway, Oliver T. Mytton, Chrissy H. Roberts, Harry Rutter, Andrea Sherriff, Martin White, Jean Adams

**Affiliations:** MRC Epidemiology Unit, Institute of Metabolic Science, University of Cambridge School of Clinical Medicine, Cambridge, UK; School of Medicine, Dentistry and Nursing, University of Glasgow, Glasgow, UK; Great Ormond Street Institute of Child Health, 30 Guilford Street, London, UK; Clinical Research Department, London School of Hygiene & Tropical Medicine, London, UK; Department of Social and Policy Sciences, University of Bath, Bath, UK

## Abstract

**Background:** Tooth extraction due to dental caries is associated with socioeconomic-deprivation and is a major reason for elective hospital admissions in England in childhood. Consumption of sugar-sweetened beverages is a risk factor for dental caries. We examined whether the soft drinks industry levy (SDIL), announced in March 2016 and implemented in April 2018, was associated with changes in incidence rates of hospital admissions for carious tooth extraction in children, 22 months post-SDIL implementation.

**Methods:** Changes in incidence rates of monthly hospital admissions for carious tooth extractions (January 2012-February 2020) in England were estimated using interrupted time series and compared with a counterfactual scenario (no SDIL announcement or implementation). Periodical changes in admissions, auto-correlation and population structure were accounted for. Estimates were calculated overall, by index of multiple deprivation (IMD) fifths and by age group (0-4, 5-9, 10-14, 15-18 years).

**Findings:** Compared to the counterfactual scenario, there was a relative reduction of 12·1% [95%CI 17·0%-7·2%] in hospital admissions for carious tooth extractions in all children (0-18 years). Children aged 0-4 and 5-9 years had relative reductions of 28·6% [95%CI 35·6-21·5] and 5·5% [95%CI 10·5%-0·5%], respectively; no change was observed for older children. Reductions were observed in children living in most IMD areas regardless of deprivation.

**Interpretation:** The UK SDIL was associated with reductions in incidence rates of childhood hospital admissions for carious tooth extractions, across most areas regardless of deprivation status and especially in younger children.

**Funding:** UK National Institute of Health and Care Research, UK Medical Research Council

**Research in Context:** *Evidence before this study:* We searched PubMed and Google Scholar for articles and reviews on the impact of sugar sweetened beverages and sugar sweetened beverage taxes on dental health, published from inception to October 15, 2022 using search terms (“sugar sweetened beverages” OR “soda” OR “soft drinks”) AND (“sugar tax” OR “sugar sweetened beverage tax” or “sugar levy”) AND (“tooth extractions” OR “tooth caries” OR “oral health” OR “dental caries”) AND (“childhood” OR “children”). Further relevant papers were found by checking reference lists of identified articles. Dental extractions due to caries are one of the most common reasons for children having an elective admission to hospital in many high income countries, including England. Identified studies suggest a strong relationship between consumption of sugar-sweetened beverages (SSBs) and the development of dental caries. The overall prevalence of caries in children has been declining for more than a decade. However, a strong social gradient exists with survey data showing children living in the most deprived areas have twice the number of decayed, missing (due to decay) and filled teeth as those living in the least deprived areas in England. Studies that have previously investigated the potential effect of SSB taxes on dental caries have mostly relied on public health modelling, with only a single empirical study based on observational data. Studies including young children (<5 years old) are particularly scarce. At present there are no studies that have examined associations between SSB taxes and changes in childhood hospital admissions for carious tooth extractions, an indicator of severe dental decay.

*Added Value of this study:* We used routinely collected nationally representative hospital episodes statistics (HES), on monthly admissions of children aged 0-18 years in England, to NHS hospitals for tooth extraction due to caries between January 2012 and February 2020. Taking account of historical trends in hospital admissions for carious tooth extraction we examined changes in these admission types in relation to the UK soft drink industry levy (SDIL), a sugar sweetened beverage (SSB) tax announced in March 2016 and implemented in April 2018. Analysis was conducted overall, and by age group and Index of Multiple Deprivation (IMD) of the child’s area of residence After accounting for existing trends, we found that the SDIL was associated with an estimated overall relative reduction of 12.1% (17.0, 7.2), in hospital admissions for carious tooth extractions in children aged 0 to 18 years. Reductions were found in children living in most IMD areas regardless of deprivation and particularly in younger children aged 0-4 and 5-9 years.

*Implications of all available evidence:* These findings add to the currently limited evidence base that SSB taxes can improve children’s dental health. These effects were seen across the spectrum of deprivation suggesting widespread population benefits and particularly in pre-school and primary school age children who have limited agency to make their own dietary decisions.

## Introduction

Dental caries (tooth decay) is the most common non-communicable disease globally^1^. In England, tooth extraction due to caries is the main reason for elective admission to hospital in children aged 5-9 years and nearly 90% of extractions in young children are due to decay.^2^ When left untreated, childhood dental caries is associated with pain, problems eating and socialising, and reduced school attendance. In England, approximately 60,000 school days are missed by children each year due to tooth extractions in hospital.^2^ The requirement for general anaesthesia, which itself is associated with distress, tiredness and bleeding^3^, is the primary reason children attend hospital for tooth extractions and is most common in young children (<4 years) and when pain is widespread.^4^

Oral health among children has been improving for more than a decade, although large inequalities still exist, with children living in the most socioeconomically deprived areas having twice the number of decayed, missing (due to decay) and filled teeth (DMFT) as those from the least deprived.^2^ Population-level interventions that have the potential to improve oral health, particularly in early-life and in deprived communities, are an important component in addressing inequalities in oral health. A multitude of risk factors for dental caries have been identified including socioeconomic factors,^5^ less-than-twice daily toothbrushing,^6^ frequent exposure to free dietary sugars^1^ and (in infants) frequent bottle feeding especially at bedtime.^7^ While the UK government have concluded that water fluoridation is a safe and cost-effective way to reduce childhood tooth decay^5^ it is not universally implemented. Furthermore fluoridation schemes on their own are not sufficient to completely prevent tooth decay meaning additional interventions are necessary.^1^

The World Health Organization (WHO) recommends added sugar should be limited to less than 10% of energy intake and that restricting sugars below 5% would provide further benefits to health, including dental health.^1^ In England, sugar sweetened beverages (SSB) are a major source of dietary added-sugars in children, accounting for around 30% of added-sugars in children 1-3 years and over 50% by late adolescence.^8^ WHO has recommended taxation of SSBs in order to reduce consumption of sugar^9^ and to date over 50 countries have implemented SSB taxes.^10^

In March 2016, the UK government announced a soft drink industry levy (SDIL) with the aim of reducing sugar intake.^11^ The two-tier tax, which was implemented in April 2018, is designed to encourage manufacturers to reformulate their drinks rather than pass the tax on to the consumer. Manufacturers of soft drinks containing ≥ 8g of sugar/100ml are subject to a levy of £ 0.24/litre and those with ≥ 5 to <8g of sugar/100ml are taxed at £ 0.18/litre. Soft drinks containing <5g/100ml sugar are not liable for the levy and 100% fruit juices, powder to make drinks, milk and milk-based drinks and drinks with 1.2% alcohol by volume or more are exempt irrespective of sugar content. Through reformulation, the UK SDIL led to large reductions in the sugar levels in soft drinks ^12^ and there was a reduction in sugar purchased from soft drinks.^13^

While the relationship between SSBs and dental caries is well-established there is limited evidence on the impacts of SSB taxes on oral health. One microsimulation study reported that an SSB tax alone was unlikely to have a significant impact on dental caries.^14^ However, other modelling studies have predicted that SSB taxes, based on a 20% tax^15–17^ or reformulation,^18^ would lead to reductions in dental caries. These studies almost exclusively focus on age-groups with permanent dentition, with some indicating the greatest benefits in children aged 15-19 years^15^ and 6-12 years, ^17^ or in children^15^ and adults^16^ from lower income households.

We are aware of only one prior empirical study on a sugar tax and dental health. That study reported that taxes on unhealthy foods and drinks in Mexico were associated with a reduction in dental caries in adults. Associations in children aged 1-12 years were lower than in adults and no associations were observed in children aged 1-5 years.^19^ However, the study did not specifically examine tooth extractions due to dental caries - an indicator of more severe caries, especially in younger children.

We used hospital episode data from England to study changes in the incidence rates of hospital admission for carious tooth extraction in children in the 22 months following the implementation of the UK SDIL (1) overall, (2) by age and 3) by area-based deprivation.

## Methods

### Data source

We used Hospital Episodes Statistics (HES) on hospital admission for dental extraction of one or more deciduous or permanent teeth due to a primary diagnosis of dental caries (International classification of diseases; ICD-10 code: K02) in children aged 0-18y in England attending a National Health Service hospital. Data included in HES were grouped and summarised by (1) index of multiple deprivation (IMD) quintile of the Lower Super Output Area (LSOA) of residence ^20^ and (2) age groups 0-4, 5-9, 10-14, 15-18 years. In HES, patient age is calculated from patient date of birth and episode start date. LSOA of residence was determined from postcode of residence. HES used the 2010 version of the Index of Multiple Deprivation (IMD) to rank LSOAs from least to most deprived and to assign records into fifths. Where postcodes were not recorded or where a link could not be made, records were excluded from the analysis (0·06%). The study period ran for 98 months from January 2012 (study month 01) to February 2020 (study month 98) and included the periods of the SDIL announcement (March 2016; study month 51) and implementation (April 2018; study month 76).

### Statistical analysis

Interrupted time series (ITS) analyses were performed to determine associations between the announcement and implementation of the SDIL and incidence rates of hospital admissions for carious tooth extractions (hereafter referred to as “hospital admissions”), in an overall model. Interaction terms revealed evidence of effect modification by age-group and IMD quintile, thus further models were run for each age and deprivation category separately (Table S1). Modelling official statistics that reported numbers annually, we used polynomial regression to estimate groupwise (i.e. age 0-4 years) population sizes in each study month. ^21^ Incidence rates of hospital admissions were then calculated by dividing the groupwise number of admissions by the respective estimated population size, multiplied by 100,000 to give an incidence rate (per 100,000 population). Time series models were based on generalised least squares (GLS).

A data driven approach, using calendar months, was used to determine periodic events associated with significant changes in hospital admissions. When calendar months were tested one-by-one, GLS models were finalised by including all the months that showed significant changes in hospital admissions. Thus, models were adjusted for the months of October, December, and March where there were statistically significant changes in incidence rates of hospital admissions.

Autocorrelation was determined using Durbin-Watson statistical tests and graphically using autocorrelation and partial-autocorrelation. For each model, an autocorrelation-moving average (ARIMA) correlation structure was selected from a plurality of possible models, with main parameters including moving average (q) and order (p) adjusted to minimise the Akaike Information Criterion (AIC) among the candidate models.

Pre-announcement trends (study months 01–51) were used to estimate counterfactual scenarios. Absolute and relative differences in the incidence rates of hospital admissions were calculated by taking the difference between observed and counterfactual values at study month 98 (February 2020). Relative change refers to the corresponding absolute change as a percentage of the counterfactual scenario value. Confidence intervals of absolute and relative differences were estimated from standard errors that were calculated using the delta method.

The main analysis included a counterfactual based on a scenario in which there was no SDIL-announcement or implementation. Whilst SDIL implementation took place in April 2018, there is evidence that drinks reformulation was underway some months before.^12^ However, since implementation marked a precise time when legal enforcement of the SDIL came into effect for eligible soft drinks, we conducted secondary analysis where counterfactual scenarios were based on pre-implementation trends (study months 01-76), rather than the pre-announcement trends in the main analysis.

Statistical analyses were conducted in R version 4.1.0. This study is registered (ISRCTN18042742) and the protocol published.^22^ Data were provided to us in an aggregated and anonymised state and ethical approval was not required for analysis of this data.

### Changes to Protocol

Several substantive changes were made to the published protocol. ^22^ Study outcomes were initially planned to be by age and deprivation and then further by gender. However, cases per 100,000 population of hospital admissions for dental extraction for dental caries each month, were deemed too small to further stratify by gender. For the same reason, our original plans to examine hospital admission by IMD tenths was revised to IMD fifths. It was planned that acute cases of asthma or appendectomy could be used as a control group however relatively unstable incidence rates by deprivation and during the study period made these conditions unsuitable as controls. The study period was ended two months earlier than planned because of the national lockdown due to the COVID-19 pandemic which began in March 2020 ^23^.

## Results

Over the 98-month study period, the mean incidence rate of hospital admissions fell from 31·0/100,000 population/month (p/m) in the preannouncement period (January 2014 – March 2018) to 28·5/100,000 p/m in the post announcement period (April 2018 – Feb 2020) (Table 1). Admissions followed a strong social gradient with incidence rates being around five times higher in those living in the most (61.6/100,000 p/m in the preannouncement period) versus least deprived areas (12.7/100,000 p/m). In terms of age, the highest incidence rate was in the 5-9 year age group (66.2/100,000 p/m in the pre-announcement period), which was approximately six times higher than in the 15-18 year age group (11.1/100,000 p/m).

**Table 1:** Mean incidence (standard deviation) of hospital admissions/100,000 population/ month for carious tooth extractions in the pre^1^ and post^2^ announcement periods of the UK SDIL, overall and by IMD quintiles and age group.

|  | Pre-announcement<sup>1</sup> | Post-announcement<sup>2</sup> |
| --- | --- | --- |
| Total population | 31.0(2.5) | 28.5(2.5) |
| Deprivation quintile: |  |  |
| IMD1 (most deprived) | 61.6(5.2) | 57.7(4.7) |
| IMD2 | 38.1(3.3) | 34.7(3.7) |
| IMD3 | 24.3(2.4) | 22.1(2.2) |
| IMD4 | 16.7(1.8) | 15.7(1.6) |
| IMD5 (least deprived) | 12.7(1.2) | 11.8(1.3) |
| Age group (years): |  |  |
| 0-4 | 21.5(2.1) | 18.3(2.3) |
| 5-9 | 66.2(5.5) | 60.7(5.4) |
| 10-14 | 21.1(2.0) | 18.8(1.7) |
| 15-18 | 11.1(1.2) | 9.4(1.1) |

<sup>1</sup> Pre-announcement period equates to the dates up to and including, January 2014 – March 2018
<sup>2</sup> Post-announcement period equates to the dates up to and including, April 2018 – Feb 2020

### Association between the UK SDIL and hospital admissions for carious tooth extractions

Unless indicated otherwise, all estimates of change in hospital admissions stated below are based on values in February 2020 at study month 98 in relation to the counterfactual scenario of no announcement or implementation of the UK SDIL.

Overall, in children aged 0-18 years, there was an absolute reduction in hospital admissions of 3·7 (95% CI: 5·2-2·2)/100,000 p/m or a relative reduction of 12·1% (95% CI: 17·0%-7·2%) compared to the counterfactual scenario (figure 1 and 2; table S2).

**Fig1.**
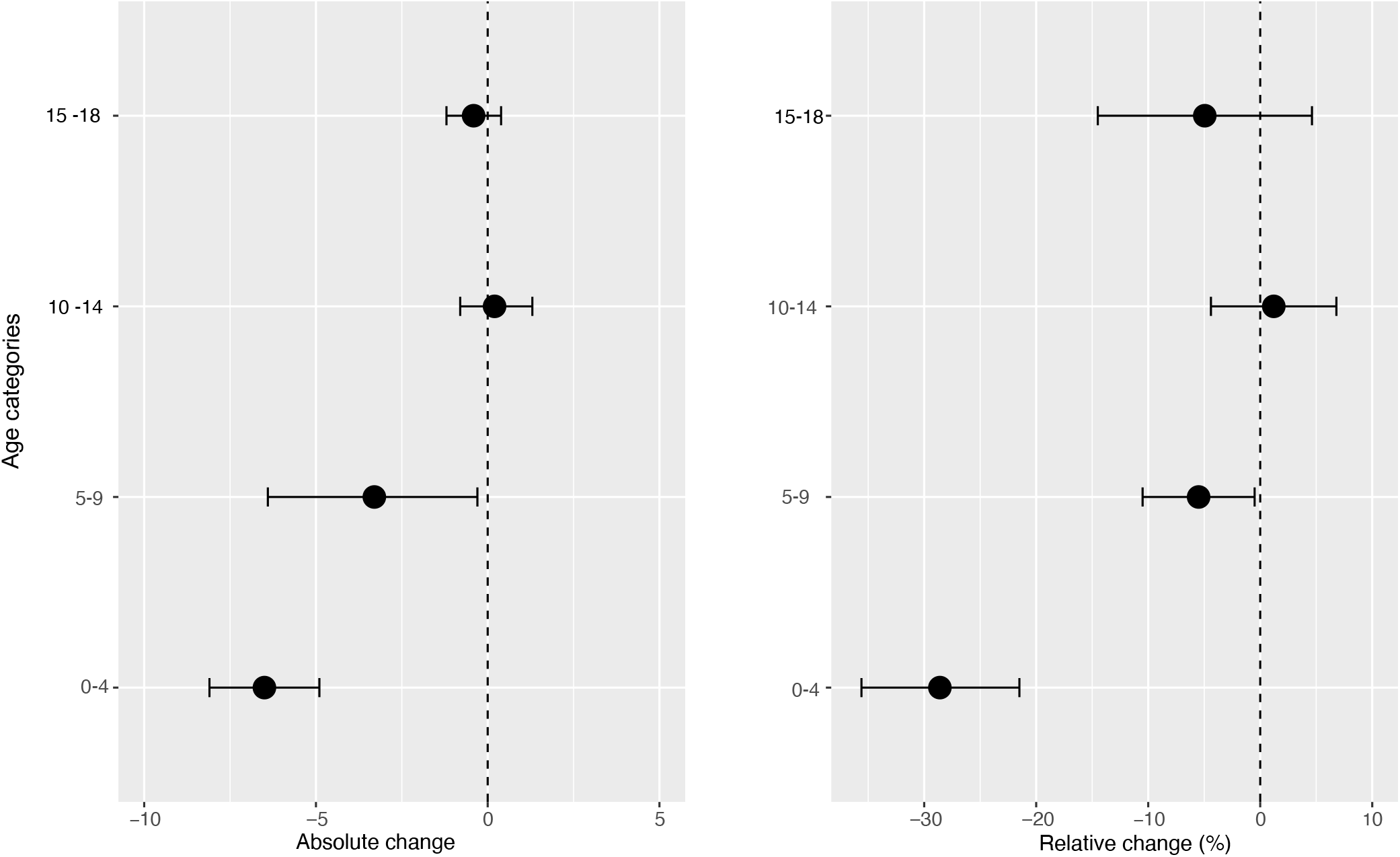
Changes in incidence/100,000 population/month of hospital admissions for carious tooth extractions (95% confidence intervals), overall and by Index of multiple deprivation (IMD) fifth at 22 months post-implementation of the UK SDIL.

**Fig2.**
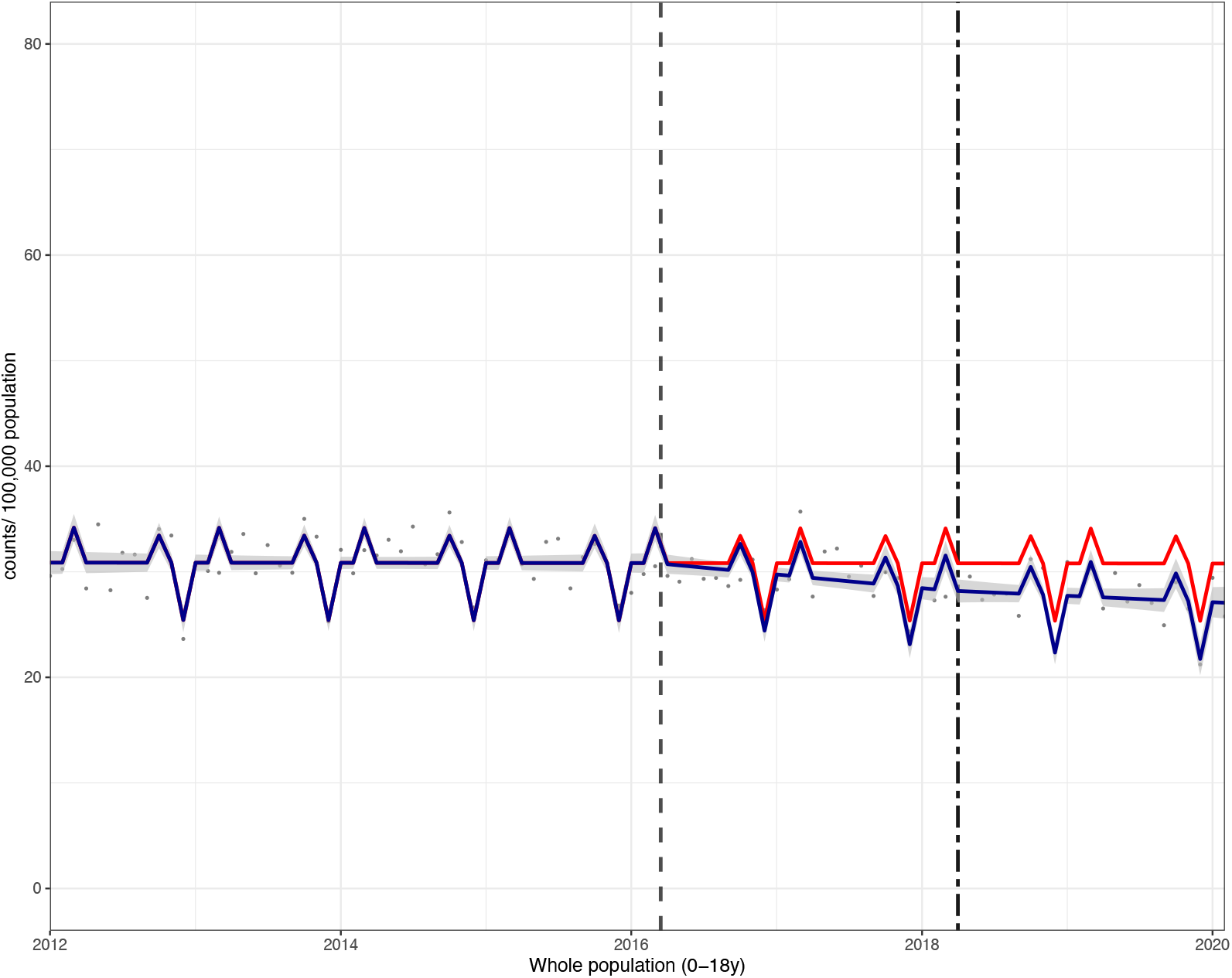
Incidence per 100,000 population per month of hospital admissions for carious tooth extractions, in children aged 0 - 18 between January 2012 and February 2020. Observed and modelled incidence of hospital admissions for carious tooth extractions is shown. Dark blue points show observed data and dark blue lines (with grey shadows) shows modelled data (and 95% confidence intervals) of incidence. The red line indicates the counterfactual line based on the pre-SDIL announcement trend (based on the announcement and implementation having not occurred). The first and second dashed vertical lines indicate the time of the SDIL announcement and implementation, respectively.

Significant reductions in hospital admissions were observed in children living in all areas regardless of deprivation, with the exception of the middle (IMD3) quintile (figures 1 and 3, table S2). For example, children living, in the second most deprived areas (IMD2) had a relative reduction in hospital admissions of 16·8% (95% CI: 22·4%-11·3%). In visualisations, a steep divergence between counterfactual and observed models is observed in IMD2 soon after the SDIL-announcement followed by notable flattening of the observed model following SDIL-implementation (figure 3).

**Fig3.**
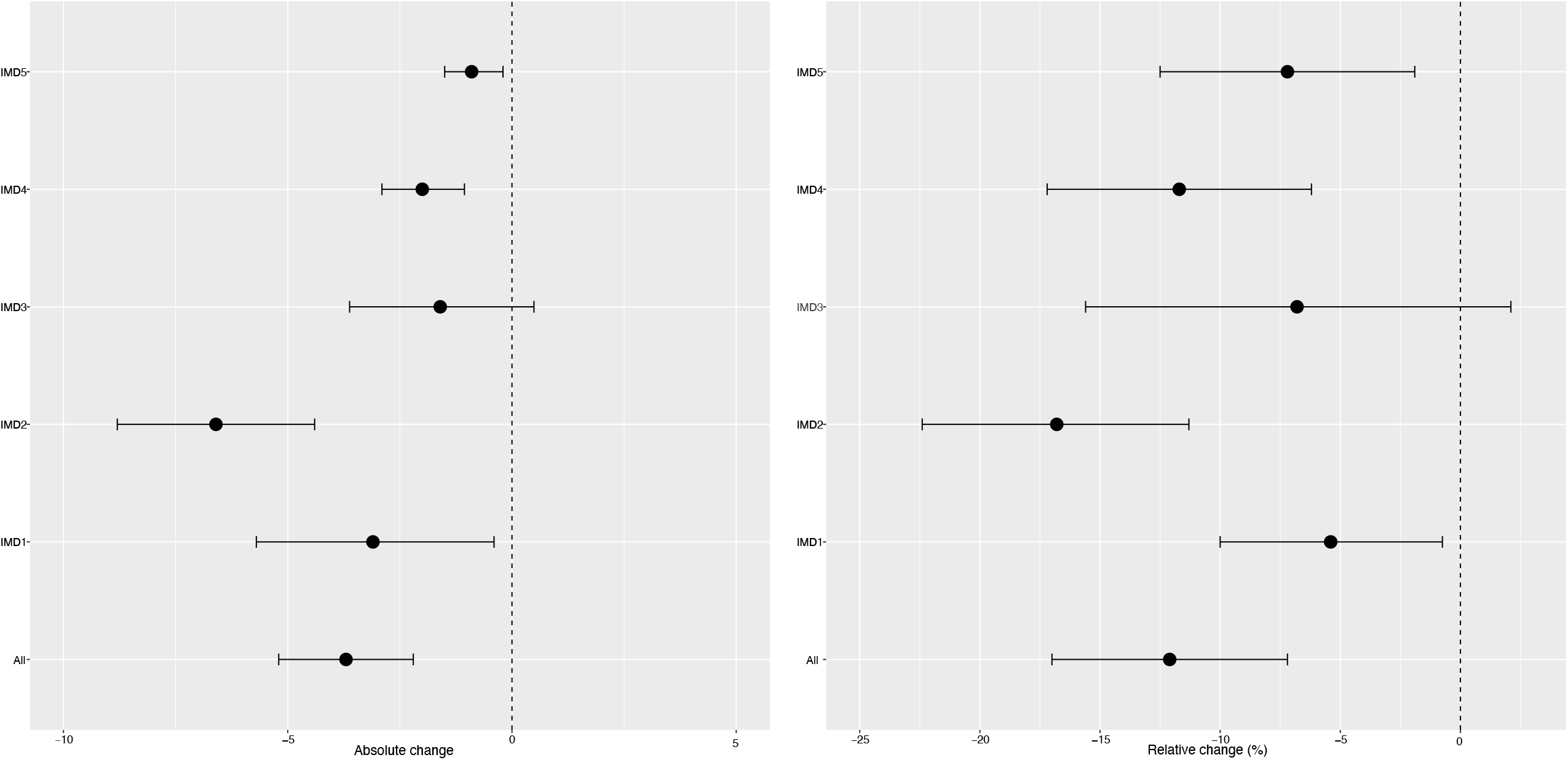
Incidence per 100,000 population per month of hospital admissions for carious tooth extractions, in children aged 0-18 between January 2012 and February 2020, by Multiple Index of Deprivation fifth. Observed and modelled incidence of hospital admissions for carious tooth extractions is shown. Dark blue points show observed data and dark blue lines (with grey shadows) shows modelled data (and 95% confidence intervals) of incidence. The red line indicates the counterfactual line based on the pre-SDIL announcement trend (based on the announcement and implementation having not occurred). The first and second dashed vertical lines indicate the time of the SDIL announcement and implementation, respectively.

The youngest children had notable reductions in hospital admissions. For example, in children aged 0-4 years a relative reduction of 28·6% (95% CI 35·6%-21·5%) were observed (figure 4 and 5, table S2). In this age group, rapid divergence between counterfactual and observed models was seen shortly after the SDIL-announcement and continued with a slight flattening of the slope following implementation. A relative reduction of 5·5% (95% CI 10·5%-0·5%), was observed for hospital admissions in children aged 5-9 years, the age-group with the greatest number of extractions (table 1). No significant changes were observed in children in age groups 10-14 and 15-18 years.

**Fig4.**
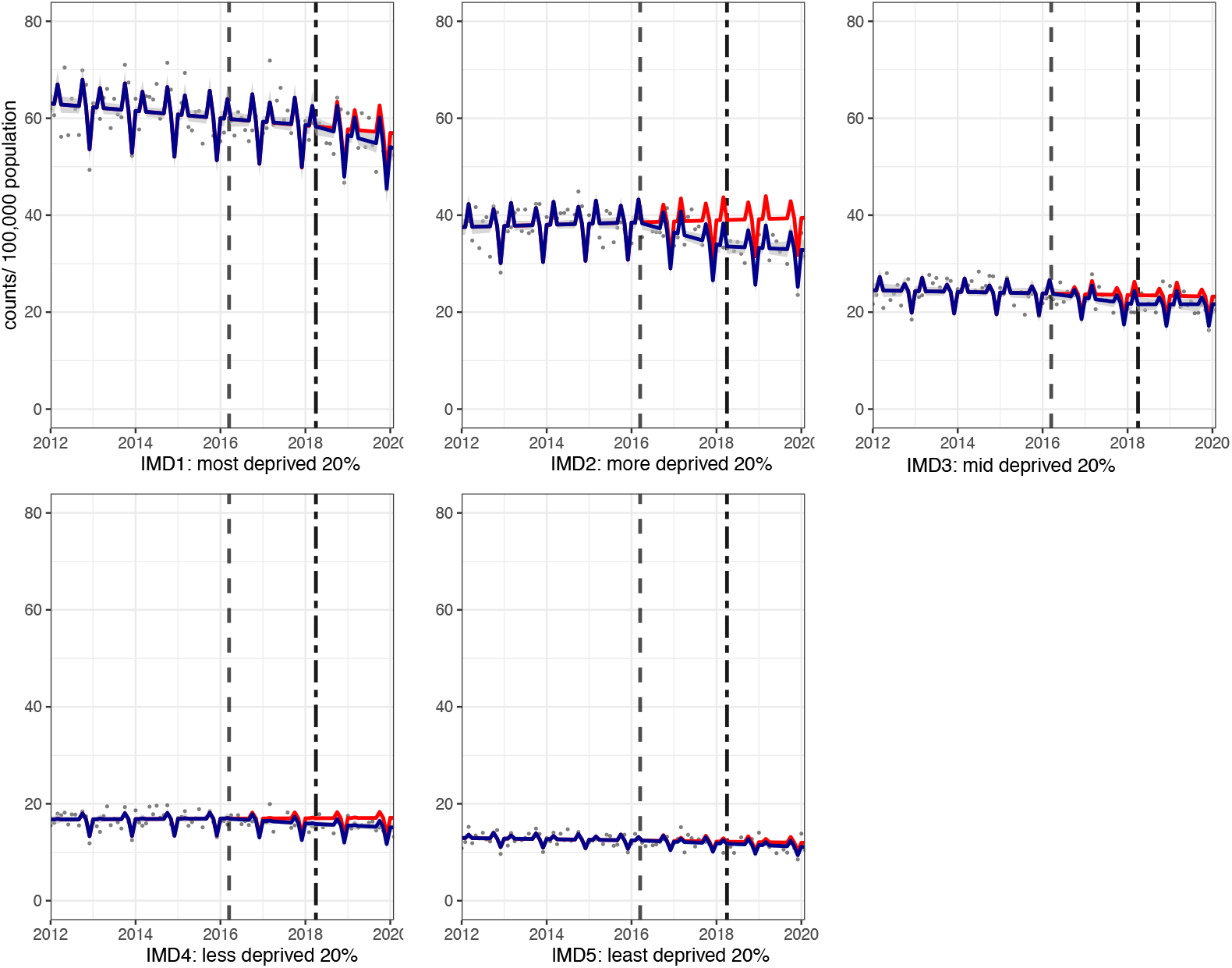
Changes in incidence/100,000 population/month of hospital admissions for carious tooth extractions (95% confidence intervals), by age group, at 22 months post-implementation of the UK SDIL.

**Fig5.**
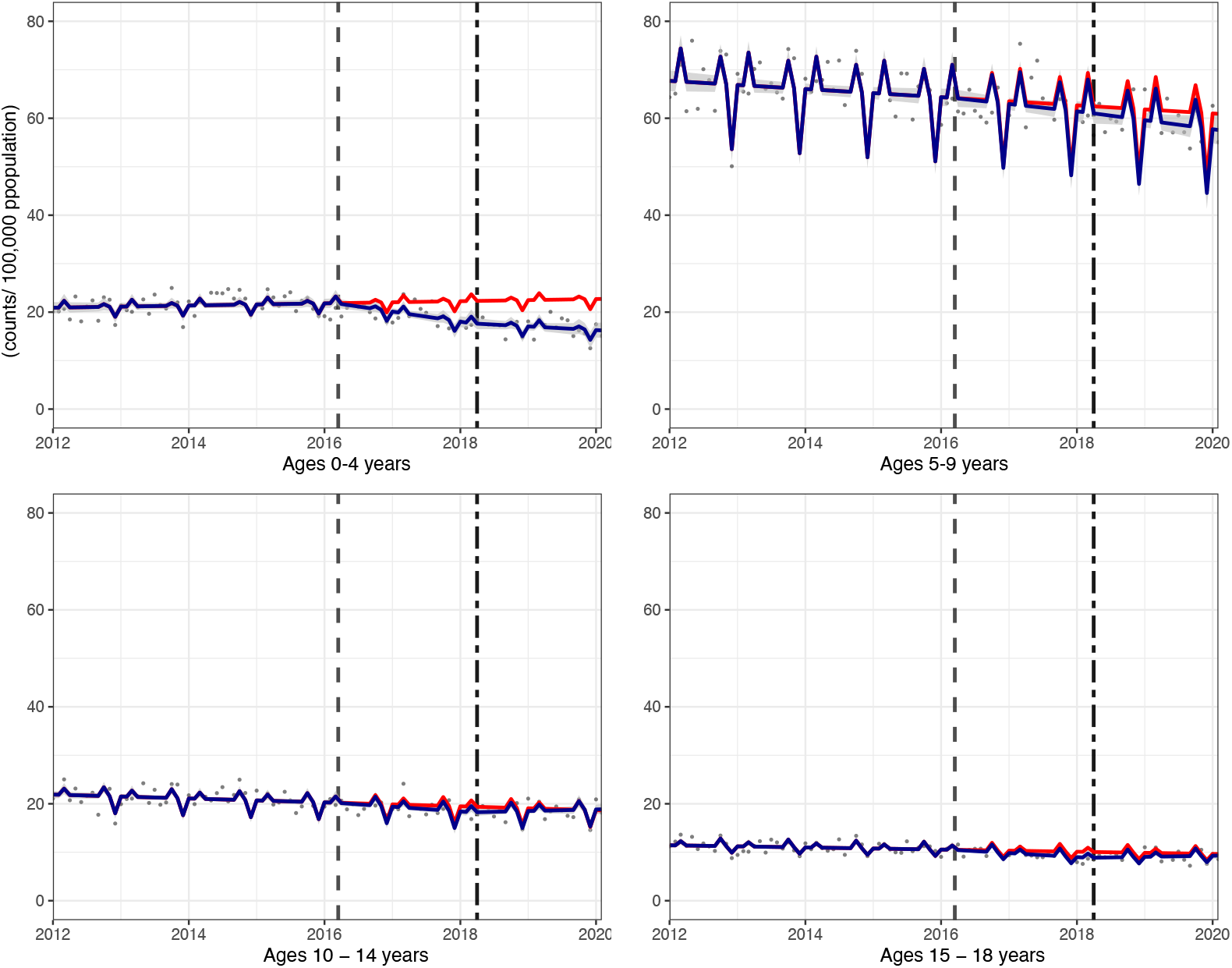
Incidence per 100,000 population per month of hospital admissions for carious tooth extractions, in children aged 0 - 18 between January 2012 and February 2020, by age-group. Observed and modelled incidence of hospital admissions for carious tooth extractions is shown. Dark blue points show observed data and dark blue lines (with grey shadows) shows modelled data (and 95% confidence intervals) of incidence. The red line indicates the counterfactual line based on the pre-SDIL announcement trend (based on the announcement and implementation having not occurred). The first and second dashed vertical lines indicate the time of the SDIL announcement and implementation, respectively.

Visualisations of the ITS models revealed notable troughs in hospital admissions in December and peaks in October and March. This may reflect periods with fewer elective surgeries due to public and school holidays, preceded or followed by catch-up periods.

In secondary analysis, we found that compared to the counterfactual scenario (of announcement but no implementation), there was no associated change in hospital admissions in children, overall (Table S3). Absolute reductions in hospital admissions of 3·3 (0·7-6·0) were observed in children living in the most deprived areas, but there were increases relative to the counterfactual in children living in IMD-2 and -3 and in all age groups except in children aged 5 to 9, where prevalence estimates were similar to the counterfactual.

## Discussion

This is the first study we are aware of to use real-world data to examine the relationship between the UK SDIL and dental health. Compared to the counterfactual of no SDIL, we found a 12·1%(95% CI 17·0%, 7·2%) relative reduction in incidence rates of hospital admissions for carious tooth extractions in children 22 months following the UK SDIL implementation. Based on a population of 12,699,899 children aged 0-18 years in England in 2020, this reduction equates to an estimated 5,638 averted cases of hospital admissions per annum.^24^ Reductions in the incidence rates of hospital admissions were observed in all deprivation groups except the middle quintile. Reductions in hospital admissions were greatest in younger children aged 0-4 and 5-9, with absolute reductions of 6·5 and 3·3/100,000 p/m, respectively. This is an important finding given that children in the 5-9 age group are the most likely to be admitted to hospital for carious tooth extractions under general anaesthesia ^25^. Incidence rates remained unchanged in older age groups (10-14 and 15-18 year).

### Strengths and limitations

This study had several strengths. First, routinely collected HES data is not subject to response bias and instead captures all NHS attendances for carious tooth extraction. The requirement for critical care hospital support for dental extractions precludes similar activity in most private UK facilities meaning we are likely to have captured almost all relevant events (with only 0.6% of admissions excluded from analysis due to non-IMD linkage). Second, the availability of area-based sociodemographic data meant that hospital admissions could be examined by IMD group. Furthermore, changing population sizes across different sociodemographic groups were accounted for over time, making our effect estimates more precise. Third, availability of monthly HES data prior to the announcement of SDIL meant that we could base our counterfactual scenarios on four years of observed data. This meant we could both detect and statistically account for predictable cyclical variations in extractions across the calendar year. Such temporally sensitive analysis could not have been conducted with the other main source of data on children’s oral health, the Children’s Dental Health Survey that takes place only every second year.^26^

A comparable control group was not available, which limits our ability to fully attribute the observed changes in hospital admissions to the SDIL. It is therefore important to consider other factors that may have influenced the outcome. To our knowledge, the only other national intervention with the potential to impact substantially on dental public health was the sugar reduction programme (2015-2020)^27^. This aimed to achieve a 20% reduction in the sugar sold in food products, but only achieved a 3.5% reduction. Nevertheless, this programme, alongside the SDIL, may have raised public awareness of sugar consumption. We are also not aware of notable changes in clinical practice during the study period. In fact, it was over a decade prior to the start point of our data (in 2000) when new regulations required that all dental general anaesthetics were carried out in hospital.^28^ We were unable to account for presence or changes in water fluoridation levels in the analysis because geo-location (LSOA) data are not made available to researchers. We note the strong evidence that associates water fluoridation schemes with reductions in dental caries^29^ but also caution that the use of water fluoridation in the UK (which is devolved to local authorities) is both geographically limited and temporally inconsistent. For example, optimal fluoridation of potable water occurred in 10.9% of LSOAs in 2014, but just 6.3% in 2016.^30^ The results of this analysis are contingent on the modelled counterfactual based on four years of data prior to the intervention and the counterfactual trajectory is subject to the usual assumptions and sources of errors that are intrinsic to any modelling approach based on counterfactuals. Finally, the IMD data provided by HES was last updated in 2010 leaving the possibility that the deprivation of areas may have changed between 2010 and the end of the study period.

### Comparison with other studies and interpretation of results

We are aware of only a few studies that have estimated the potential impacts of sugar taxes on dental caries in very young children (<5 years) using either health impact modelling^18^ or empirical data.^19^ In contrast to our findings of greatest effects in younger groups, a modelling study predicted the SDIL would have greatest relative reductions in DMFT in English children aged 11-18 years as they had the highest baseline SSB consumption.^18^ That study, as well as using DMFT rather than incident hospital admissions for tooth extractions, assumed a uniform effect of sugar (and SSBs) on children’s teeth. Our contrasting findings may be explained by the intrinsic compositional differences between deciduous and permanent teeth.^18^ Deciduous teeth have a thinner enamel covering than permanent teeth which typically begin to erupt in children after 6 years of age. This means that the relationship between SSB consumption and caries may be stronger in younger than in older children. Thus, the timeline of permanent teeth eruption is an important factor to account for when comparing the impact of oral health interventions on children of different ages.^31^

Age-related differences in feeding practices may also be an important reason, not factored into the previous modelling study, that could explain why younger children may appear to disproportionately benefit from SDIL-prompted reformulation.^18^ Expert opinion advises that use of infant bottles and sippy cups, especially those containing SSBs, should be avoided in the hour before bedtime because it increases the risk of dental caries due to a dramatic reduction in saliva production during sleep; ultimately leading to slow removal of dietary free sugars and in turn a low pH and a prolonged period of demineralisation of the teeth.^33^ One study found that approximately 50% of four-year-olds use a sippy cup and 11% continue to use an infant bottle.^32^ The use of these drinking vessels is likely to be rare in older children and thus in the context of SSBs and SDIL, infant bottles and sippy cups may therefore represent a unique risk factor in younger age groups that warrants further investigation.

In contrast to our findings, an empirical study examining taxes on unhealthy food and drink in Mexico reported reductions in dental caries in older children and adults but not children aged 1-5 years^19^. Structural differences in the Mexican and UK taxes may explain these differences. In Mexico, the volume-based tax was fully passed on to the consumer in the form of increasing retail prices^34^ but there was no incentivisation for manufacturers to reformulate and reduce the sugar content of SSBs. In contrast, the UK SDIL was based on sugar-density and was designed to encourage manufacturers to reformulate soft drinks, which they did.^12^ Furthermore, the Mexico study examined incident dental caries rather than the more severe carious dental extractions, precluding a direct comparison.

We observed reduced incidence rates of tooth extractions in children from all IMD fifths, except the middle group (IMD-3). In contrast, a previous modelling study suggested SSB taxes would lead to greatest reductions in caries in the lowest income groups as they have the highest baseline SSB consumption. ^15^ One potential reason why we did not find a clear trend in effect by IMD is that water fluoridation is more common in the most deprived areas in England. Nearly one fifth (18.7%) of the population of England living in areas with fluoridated drinking water, live in the most deprived tenth of areas.^35^ This additional protective factor was not considered in the modelling study. ^15^

Our finding of an overall reduction in hospital admissions associated with the SDIL was not replicated in secondary analysis where the interruption point was moved to the date of SDIL implementation. This suggests that the biggest benefits to oral health occurred in the period between the SDIL announcement and implementation, when most reformulation took place.^12^

## Conclusion

Using administrative data on incidence rates of hospital admission for carious tooth extractions, we found the announcement of the UK SDIL was associated with improvements in children’s oral health. These benefits were observed in children living in most areas regardless of deprivation and particularly in the youngest children (<9 years). This study provides evidence of possible benefits to children’s health from the UK SDIL beyond obesity which it was initially developed to address.

## Data Availability

Data was acquired through a data sharing agreement with NHS digital for which conditions of use apply. Requests for data must be made directly to NHS digital and cannot be granted by the authors

https://digital.nhs.uk/data-and-information/data-tools-and-services/data-services/hospital-episode-statistics

## Figure legends

**Table S1:** Coefficients of interaction between study phases, IMD and age-groups.

|  | Estimate<sup>1</sup> | Standard error | P-value |
| --- | --- | --- | --- |
| **IMD fifths** |  |  |  |
| Phase: Pre-announcement |  |  |  |
| IMD1 (least deprived) | ref | ref | ref |
| IMD2 | 0.01 | 0.03 | 0.67 |
| IMD3 | -0.01 | 0.03 | 0.67 |
| IMD4 | 0.02 | 0.03 | 0.44 |
| IMD5 (most deprived) | -0.06 | 0.08 | 0.03* |
| Phase: Post-announcement - pre-implementation |  |  |  |
| IMD1 (least deprived) | ref | ref | ref |
| IMD2 | -0.03 | 0.08 | 0.71 |
| IMD3 | -0.04 | 0.08 | 0.60 |
| IMD4 | -0.19 | 0.08 | 0.01* |
| IMD5 (most deprived) | 0.04 | 0.08 | 0.61 |
| Post-implementation phase |  |  |  |
| IMD1 (least deprived) | ref | ref | ref |
| IMD2 | 0.01 | 0.12 | 0.92 |
| IMD3 | 0.06 | 0.12 | 0.64 |
| IMD4 | 0.13 | 0.12 | 0.29 |
| IMD5 (most deprived) | -0.18 | 0.12 | 0.14 |
| **Age groups** |  |  |  |
| Phase: Pre-announcement |  |  |  |
| 0-4 years | ref | ref | ref |
| 5-9 years | -0.10 | 0.03 | 0.0007* |
| 10-14 years | -0.05 | 0.03 | 0.07 |
| 15-18 years | -0.04 | 0.03 | 0.21 |
| Phase: Post-announcement - pre-implementation |  |  |  |
| 0-4 years | ref | ref | ref |
| 5-9 years | 0.17 | 0.08 | 0.04* |
| 10-14 years | 0.14 | 0.08 | 0.07 |
| 15-18 years | 0.14 | 0.08 | 0.09 |
| Phase: Post-implementation |  |  |  |
| 0-4 years | ref | ref | ref |
| 5-9 years | -0.2 | 0.13 | 0.13 |
| 10-14 years | <0.01 | 0.13 | 1.00 |
| 15-18 years | -0.01 | 0.13 | 0.93 |

<sup>1</sup>Change in incidence rate (n/100,000 population/month)

**Table S2:** Changes^1^ in incidence/100,000 population/month of hospital admissions for carious tooth extractions (95% confidence intervals), overall and by Index of multiple deprivation (IMD) fifth and age group at 22 months post-implementation of the UK SDIL.

|  | Absolute change | Relative change (%) |
| --- | --- | --- |
| Total population | **-3.7(-2.2, -5.2)** | **-12.1(-17.0, -7.2)** |
| Deprivation fifths |  |  |
| IMD1 (most deprived) | **-3.1(-0.4, -5.7)** | **-5.4(-10.0, -0.75)** |
| IMD2 | **-6.6(-4.4, -8.8)** | **-16.8(-22.4, -11.3)** |
| IMD3 | -1.6(0.49, -3.62) | -6.8(-15.6, 2.1) |
| IMD4 | **-2.0(-1.06, -2.9)** | **-11.7(-17.2, -6.2)** |
| IMD5 (least deprived) | **-0.9(-0.2, -1.5)** | **-7.2(-12.5, -1.9)** |
| Age Group: |  |  |
| 0-4 | **-6.5(-4.9, -8.1)** | **-28.6(-35.6, -21.5)** |
| 5-9 | **-3.3(-0.3, -6.4)** | **-5.5(-10.5, -0.5)** |
| 10-14 | 0.2(1.3, -0.8) | 1.21(-4.4, 6.8) |
| 15-18 | -0.41(0.39, -1.2) | -4.95(-14.5, 4.62) |

<sup>1</sup>Absolute and relative changes compared to the counterfactual scenario, which is based on a scenario of no SDIL announcement and no implementation

**Table S3:**
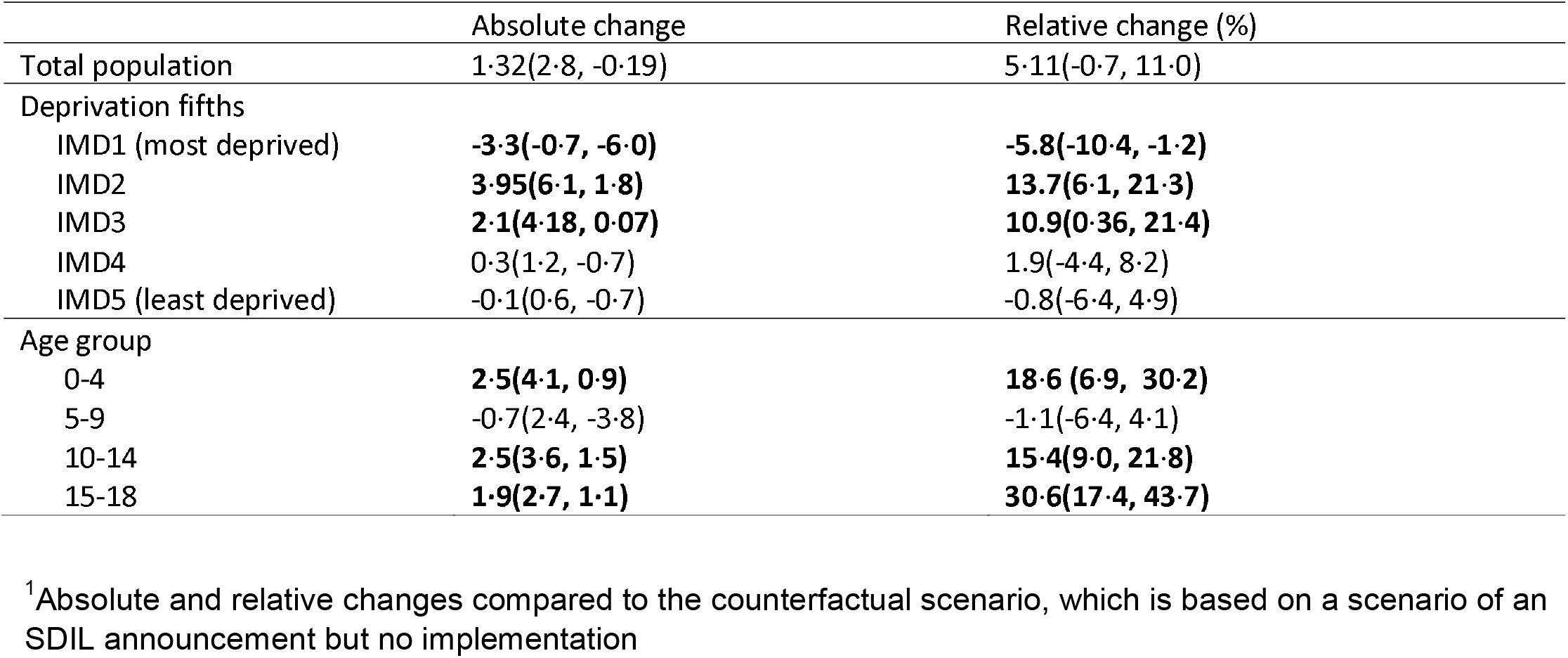
Changes^1^ in incidence /100,000 population/month of hospital admissions for carious tooth extractions (95% confidence intervals), overall and by Index of multiple deprivation (IMD) fifth and age group at 22 months post SDIL implementation (secondary analysis).

|  | Absolute change | Relative change (%) |
| --- | --- | --- |
| Total population | 1.32(2.8, -0.19) | 5.11(-0.7, 11.0) |
| Deprivation fifths |  |  |
| IMD1 (most deprived) | **-3.3(-0.7, -6.0)** | **-5.8(-10.4, -1.2)** |
| IMD2 | **3.95(6.1, 1.8)** | **13.7(6.1, 21.3)** |
| IMD3 | **2.1(4.18, 0.07)** | **10.9(0.36, 21.4)** |
| IMD4 | 0.3(1.2, -0.7) | 1.9(-4.4, 8.2) |
| IMD5 (least deprived) | -0.1(0.6, -0.7) | -0.8(-6.4, 4.9) |
| Age group |  |  |
| 0-4 | **2.5(4.1, 0.9)** | **18.6(6.9, 30.2)** |
| 5-9 | -0.7(2.4, -3.8) | -1.1(-6.4, 4.1) |
| 10-14 | **2.5(3.6, 1.5)** | **15.4(9.0, 21.8)** |
| 15-18 | **1.9(2.7, 1.1)** | **30.6(17.4, 43.7)** |

<sup>1</sup>Absolute and relative changes compared to the counterfactual scenario, which is based on a scenario of an SDIL announcement but no implementation

